# Real-World Outcomes and Predictors of Accelerated rTMS Treatment Response for Treatment-Resistant Depression

**DOI:** 10.1101/2024.05.05.24306898

**Authors:** Danielle D. DeSouza, Erica Nakano, Vivian Hoang, Kim Hoang, David Ling, Owen Muir, Nathan Meng, Noah DeGaetano, Shan H. Siddiqi, David Carreon

**Author notes:** **Corresponding Author:** David Carreon, MD, 877 W. Fremont Ave, Suite N-3, Sunnyvale, CA, 94087. indicates co-last authorship (Shan H. Siddiqi, David Carreon).

## Abstract

**Introduction:** Accelerated repetitive transcranial magnetic stimulation (rTMS) is a high-dose precision-targeted treatment that can induce rapid remission from depressive symptoms with only five days of treatment. However, real-world outcomes and the various demographic, clinical, and treatment variables that influence its effectiveness are not fully known.

**Objective:** To determine real-world outcomes of accelerated rTMS treatment for treatment-resistant depression and identify predictors of treatment response.

**Methods:** This retrospective analysis included 226 individuals treated in an outpatient clinic. Demographics, clinical history, and accelerated rTMS treatment parameters were extracted from medical records. Outcomes were evaluated within 1-month of treatment and general linear models examined predictors of treatment response in various subgroups.

**Results:** Accelerated rTMS demonstrated substantial response rates within the first month (66% with neuronavigation; 63% without neuronavigation). Response rate was independent of the number of antidepressant failures, prior ECT, or prior TMS, illustrating its utility for patients with a high degree of treatment resistance. Significant predictors of positive treatment response included resting-state functional magnetic resonance imaging (fMRI)-guided targets and female gender.

**Discussion:** Accelerated rTMS is effective for treatment-resistant depression in an outpatient clinical setting. fMRI-guided targeting significantly improved outcomes, suggesting that it provides added benefit over conventional targeting. Additionally, female patients showed better responses, in line with previous literature. These findings highlight the value of personalized approaches in clinical practice and warrant further investigation in larger clinical trials.

## INTRODUCTION

Accelerated repetitive transcranial magnetic stimulation (rTMS) represents a promising shift in the acute management of depressive disorders, aiming to address the delay in symptom improvement experienced with traditional pharmacotherapy and conventional once-daily rTMS treatments^1^.

Innovations to this approach include integrating resting-state functional magnetic resonance imaging (fMRI) to personalize the targeting of the left dorsolateral prefrontal cortex (DLPFC) based on individual functional connectivity to the subgenual anterior cingulate cortex (sgACC) ^2–4^. This approach can also be used to stimulate distinct brain circuit targets that can improve ‘dysphoric’ or ‘anxiosomatic’ symptom clusters^5^.

Image-guided targeting differs from conventional techniques, which use scalp measurements to identify a TMS target and do not account for interindividual variability in brain structure and function, potentially contributing to inconsistent treatment outcomes^6^.

Variation in treatment outcome may also be related to other clinical, demographic, or treatment factors, such as comorbidities, age, sex, or the number of treatment sessions, although these associations have been inconsistent in previous studies^7^. The goal of this study is to describe real-world treatment outcomes for individuals receiving accelerated rTMS for treatment-resistant depression symptoms, and evaluate potential predictors of response. Assessing real-world outcomes provides a broader perspective on the effectiveness and utility of accelerated rTMS outside of controlled clinical trials, and reflects a more diverse patient population under varied treatment protocols. This comprehensive view can provide insight into the factors contributing to accelerated rTMS treatment success and enhance our understanding of its utility in routine clinical practice.

## METHODS

This retrospective analysis included 226 patients with depressive symptoms who received accelerated rTMS treatment at Acacia Clinics, an outpatient clinic focused on accelerated TMS in Sunnyvale, California, from July 2018 to October 2023. Patients with a primary diagnosis of major depressive disorder (MDD) or bipolar disorder (BPD), with or without comorbidities, were included. Pearl IRB granted exemption to this study as secondary research uses of data and no Institutional Review Board (IRB) approved informed consent was required.

Physicians clinically evaluated patients and discussed the state of the evidence for neuronavigation vs non-navigation. Known risks and benefits, patient preference, and cost were all considered; neuronavigation costs were a common deciding factor for patients.

Demographic details, clinical history, and accelerated rTMS treatment parameters were extracted from medical records through a retrospective chart review. To assess treatment outcomes, patients completed the Patient Health Questionnaire-9 (PHQ-9) prior to treatment and within 1-month following treatment. The PHQ-9 is a nine-item, self-report depression scale. Scores for each question range from 0 to 3 (“not at all” to “nearly every day”). Total scores of 0-5, 5-10, 10-15, and 15-20 indicate mild, moderate, moderately-severe, and severe depression, respectively^8^. Remission is defined as a post-treatment score of less than 5^9^, and response is defined by a ≥50% decrease in the baseline PHQ-9 total score.

In cases where the post-treatment PHQ-9 score was not available, outcomes within 1-month of treatment were evaluated by a psychiatrist using the Clinical Global Impressions-Improvement (CGI-I) Scale. The CGI-I is scored based on the comparison of symptom presentation from baseline to the present evaluation, taking into account both reported and observed patient symptoms, behavior, and function^10^. CGI-I scores range from 1 (very much improved) to 7 (very much worse), with scores of 1, 2, and ≥ 3 indicating remission, response, and nonresponse, respectively.

Accelerated rTMS treatments were administered using the MagVenture MagPro R30 system (MagVenture A/S, Denmark) equipped with a MagVenture Cool-B65 coil. Most patients received 10 stimulation sessions per day spaced by 50-minute inter-session intervals.^4,11^ Treatments were delivered using intermittent theta-burst stimulation (iTBS) (1800 pulses per session); however, in a few cases high-frequency stimulation (10-20 Hz), low-frequency stimulation (1 Hz), or continuous theta-burst stimulation (cTBS) protocols were used.

Treatments were prescribed as 80-120% of the resting state motor threshold, where motor threshold was determined as the minimum amplitude necessary to evoke a hand movement response 50% of the time^12^. For most patients, a single left DLPFC target was stimulated. In some cases, right-sided treatment, bilateral treatment, or target switching part-way through the treatment course was done based on clinical factors.

For patients with resting-state fMRI-derived targets, MRI scanning sessions were completed prior to treatment. Scans were acquired at different imaging facilities based on availability. In all cases, resting-state fMRI scans were at least 20-minutes in duration. High-resolution T1-weighted anatomical scans were also acquired. Prior to resting-state connectivity analysis, scans were preprocessed as described previously^13^. Briefly, steps included spatial alignment, atlas registration, ART-based motion censoring (framewise displacement threshold of 0.5 mm), removal of respiratory artifacts, global signal regression, temporal filtering, and spatial smoothing, using SPM12 and the CONN toolbox. Individual-level precision functional mapping of brian networks was conducted using in-house scripts based on the Infomap algorithm for modular community detection. Seed-based connectivity was also used to generate connectivity maps for specific regions of interest.

Targets were then chosen manually by a neuropsychiatrist with expertise in MRI-guided TMS (SHS) based on these connectivity maps as well as patient-specific symptom profiles. The most commonly used resting-state fMRI-derived targets included the region of left DLPFC with the greatest anti-correlation to the sgACC in the majority of patients (examples in Fig. 1a-1b). For some patients, the right ‘anxiosomatic’ target^5^ was chosen based on individualized precision functional mapping of the default mode network, which overlaps closely with the anxiosomatic circuit. The right DLPFC with the greatest anti-correlation to the sgACC was also used in some cases. The Localite TMS Navigator system was used to maintain target accuracy and track motion during treatment sessions, ensuring precise alignment throughout.

**Figure 1:**
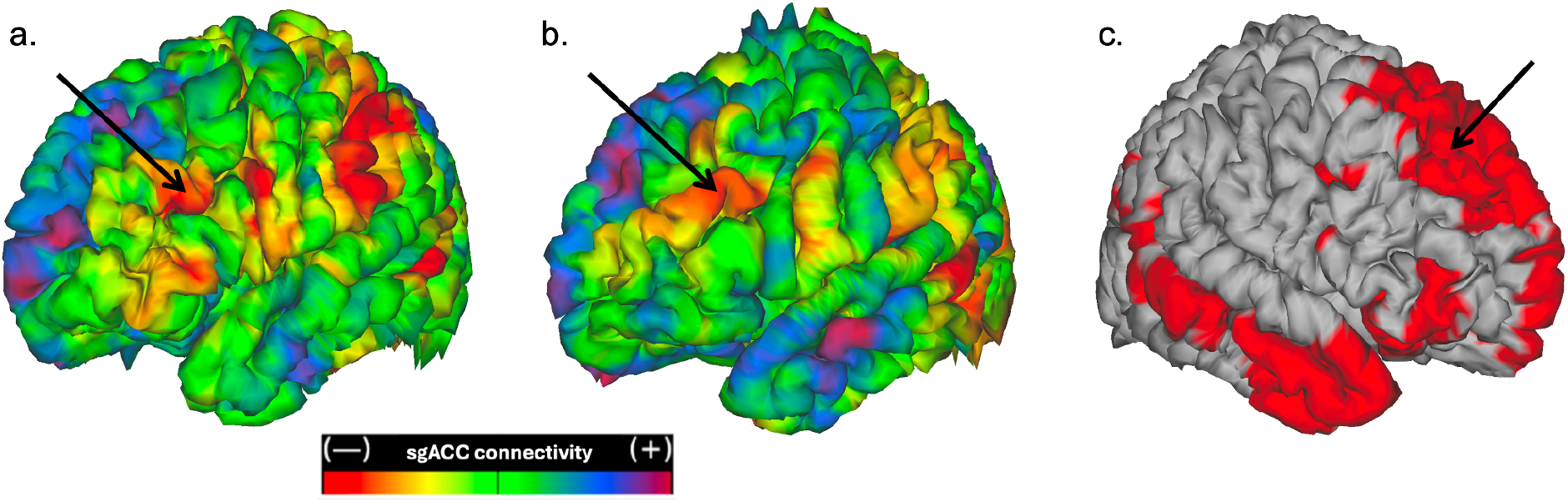
Representative resting-state fMRI connectivity-based targets. For most participants who received resting-state fMRI-based targets, individualized left DLPFC targets were generated based on the greatest anti-correlation to the sgACC (panels a-b show two representative examples). Regions outside the DLPFC were not considered due to inadequate evidence for safety or efficacy of these targets. The right ‘anxiosomatic’ target^5^ was also used for some treatments (panel c). Black arrows represent the approximate peak stimulation site.

The other approach used for targeting the left DLPFC was the Beam F3 method, which approximates the F3 location on the 10-20 system for standardized placement of electroencephalogram (EEG) electrodes^14^. This location can be determined without EEG equipment using three scalp measurements. It accounts for individual variability in head size but does not require neuroimaging guidance for target localization^15^. Beam F4 corresponds to the right DLPFC and can be safely used during bilateral stimulation sessions where left-sided stimulation is followed by right-sided stimulation or as an alternative treatment target to left-sided stimulation; because of its effect treating mania, it was often added in patients with BPD to prevent emergence of mania^16^.

Treatment outcomes were summarized for all patients and in subgroups of interest based on PHQ-9 scores or CGI-I scores as described above. To examine predictors of treatment outcome, the analysis was restricted to a subset of patients who received only left-sided DLPFC treatment, enabling comparability with existing literature and minimizing confounds related to varying stimulation sites. A general linear model (GLM) was used to identify significant predictors of treatment outcome using MATLAB R2023b (MathWorks, Natick MA). Predictor variables included target type (fMRI-guided or not), age, sex, presence of comorbidities beyond MDD, baseline PHQ-9 score, number of stimulation sessions per day, total number of stimulation sessions, past TMS treatment, past electroconvulsive therapy (ECT) treatment, and number of failed antidepressant trials. The clinical variables examining past treatments and failed antidepressant trials were included as indicators of treatment-resistance.

We had a wide range of patients and tailored the treatments to the symptoms. We had a subgroup of patients that had predominantly depressive symptoms which were similar enough to one another that we could more carefully isolate the effect of neuronavigation. All patients with predominantly depressive symptoms (i.e. they did not have a substantial degree of anxiety, OCD, etc) were prescribed a left DLPFC target, either at non-neuronavigated Beam F3 or at the neuro-navigated dysphoric target. We ran a GLM to address the question of whether neuronavigation showed benefit beyond TMS acceleration.

## RESULTS

Demographic, clinical history, and treatment variables are summarized in Table 1 for the 226 patients included in the study. To evaluate treatment outcomes, PHQ-9 scores were used. In cases where PHQ-9 scores were not available (n= 47), CGI-I scores were used instead.

**Table 1:** Demographic, Clinical, and Treatment Variables for all Patients.

|  |  |  |
| --- | --- | --- |
| Demographic Variables | Age | 45.6 +/- 19.4 |
|  | Gender | Man: 117 (52%)<br>Woman: 105 (46.5%)<br>Transgender: 1 (0.5%)<br>Not reported: 3 (1%) |
| Clinical Variables | Primary Diagnosis | MDD (no comorbidities): 84 (37.2%)<br>MDD (with comorbidities): 123 (54.4%)<br>BPD (no comorbidities): 12 (5.3%)<br>BPD (with comorbidities): 7 (3.1%) |
|  | Comorbidities (Among All) | GAD: 50 (22.1%)<br>PTSD: 13 (5.7%)<br>OCD: 22 (9.7%)<br>ADHD: 10 (4.4%)<br>Other anxiety: 40 (17.7%)<br>Other: 69 (30.5%) |
|  | Past ECT (n= 199) | Yes: 25 (13%)<br>No: 174 (87%) |
|  | ECT outcome (n= 25) | Effective: 14 (56%)<br>Ineffective: 7 (28%)<br>Unknown: 4 (16%) |
|  | Past TMS (n= 199) | Yes: 54 (27%)<br>No: 118 (59%)<br>Unknown: 27 (14%) |
|  | Benzodiazepines (n=205) | Yes: 35 (17%)<br>No: 170 (83%) |
|  | # of Previous Antidepressants | 5.8 +/- 5.6 |
|  | Baseline PHQ-9 | 16.7 +/- 5.4 |
| Treatment Variables | # of Accelerated Sessions | 50.4 +/- 14.0 |
|  | Mean # of Sessions/ Day | 8.8 +/- 2.5 |
|  | Target | Neuronav Targets (n=153) |
|  |  | L-DLPFC: 86 (56%)<br>R-Anxiosomatic: 24 (16%)<br>Bilateral: 10 (6.5%)<br>L -> R: 9 (6%)<br>R -> L: 10 (6.5%)<br>L->Bilateral: 4 (2.5%)<br>Other: 9 (6.5%)<br>Non-Neuronav Targets (n=73)<br>L-DLPFC (Beam F3): 26 (35.6%)<br>R-DLPFC (Beam F4): 1 (1.4%)<br>Bilateral: 22 (30.1%)<br>L -> R: 1 (1.4%)<br>R -> L: 1 (1.4%)<br>L -> Bilateral: 8 (10.9%)<br>Other: 14 (19.2%) |

**Abbreviations:** MDD: Major Depressive Disorder, BPD: Bipolar Disorder, GAD: Generalized Anxiety Disorder, PTSD: Post-Traumatic Stress Disorder, OCD: Obsessive-Compulsive Disorder, ADHD: Attention Deficit Hyperactivity Disorder, ECT: Electroconvulsive Therapy, TMS: Transcranial Magnetic Stimulation, PHQ-9: Patient Health Questionnaire-9, L-DLPFC: Left Dorsolateral Prefrontal Cortex, R-DLPFC: Right Dorsolateral Prefrontal Cortex, R-Anxiosomatic: Right 'Anxiosomatic' target, L: left-sided stimulation, R: right-sided stimulation. Arrows (->) indicate that a target was switched part-way through the treatment course. Bilateral indicates that left and right targets were sequentially stimulated within a treatment session (not simultaneously). For the Neuronav (neuronavigated, fMRI-Based Targets), patients who had hybrid treatments, meaning a portion of their treatment included a non-neuronavigated (non-neuronav, Beam Target) (n= 14), are included. Data in the table represent mean +/- standard deviation (SD) or number (proportion, %) as appropriate. Sample size is noted when data for the full cohort were not available.

**Table 2:**
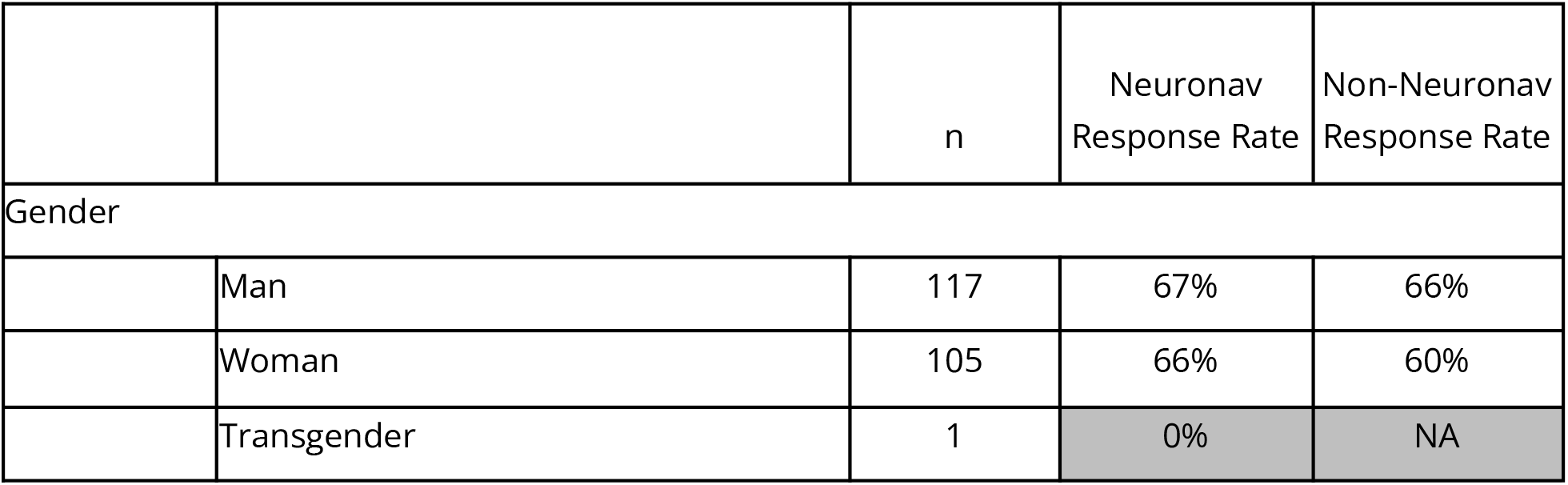
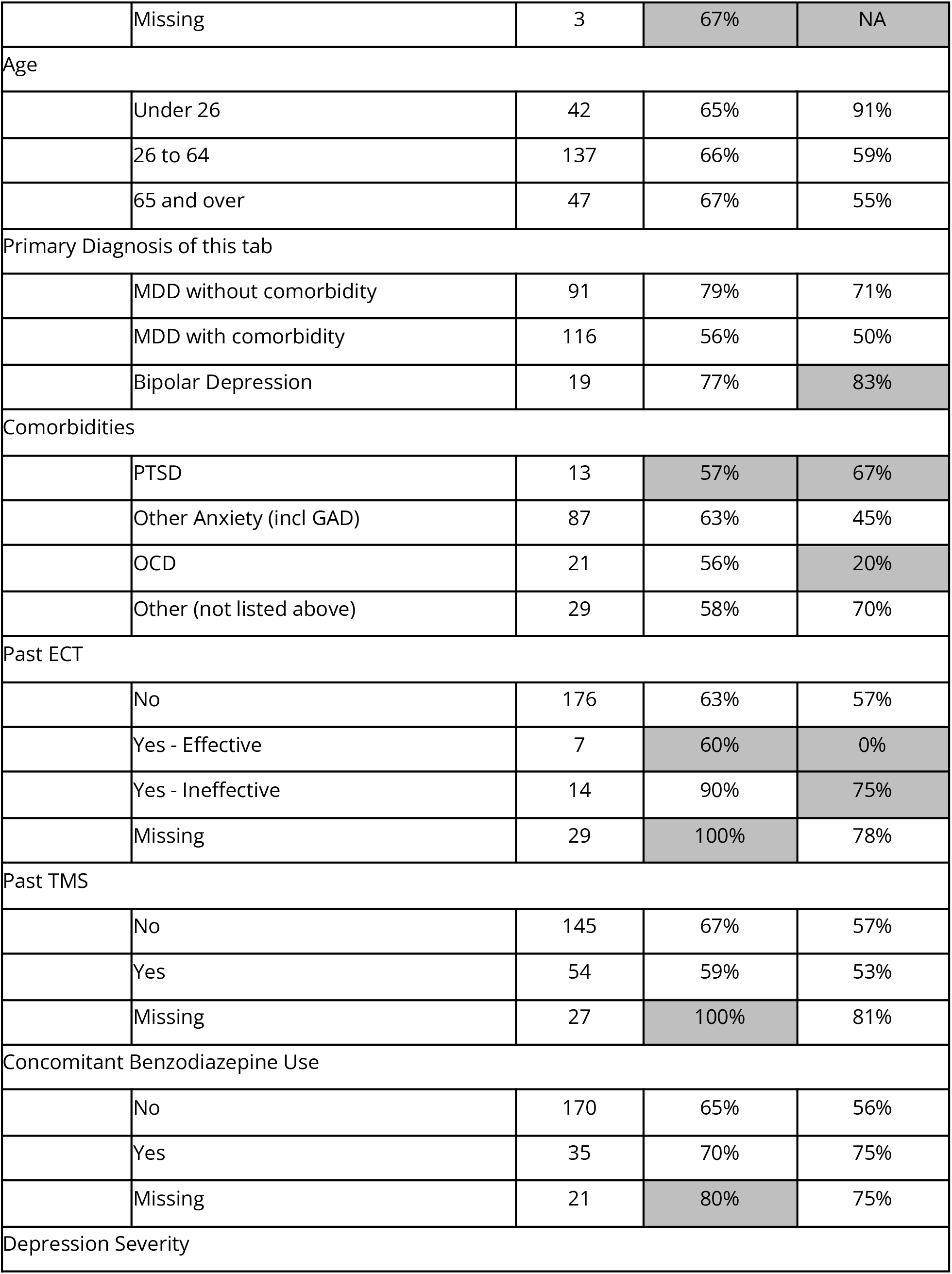
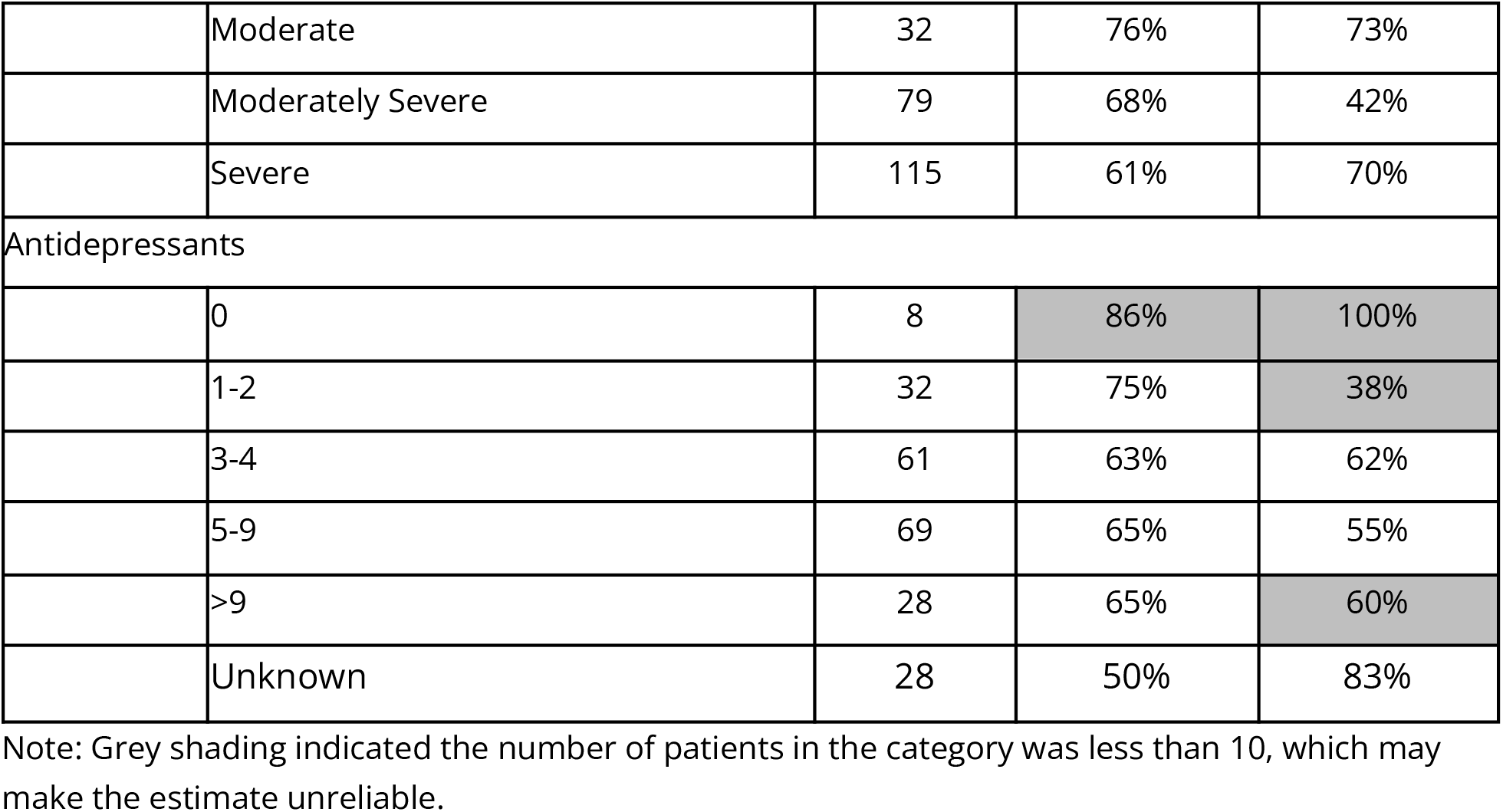
Response Rates.

The response rates were grossly similar across categories, including patients with bipolar depression, patients with comorbid conditions, patients who trialed ECT, patients who trialed TMS, patients on concomitant benzodiazepines, and patients with a high past antidepressant count.

To assess predictors of treatment outcome, a GLM was conducted on a subset of 111 patients who received treatment to the left DLPFC. The average age of this group was 45.1 years (SD= 19.2) and included 66 men, 43 women, and 1 transgender individual (1 person did not respond). Significant predictors of response included targeting type, with resting-state fMRI-based targets yielding better outcomes (Estimate= 0.358, SE= 0.148, t= 2.412, *p*= 0.018), resulting in an odds ratio of 1.43 for improved treatment response. Additionally, gender was a significant factor, with women experiencing more favorable outcomes (Estimate= -0.263, SE= 0.106, t= -2.471, *p*= 0.016). Of note, patients receiving neuronavigation appeared to have a greater degree of treatment resistance (mean failed antidepressants = 6.1 ± 4.9) relative to those who did not receive neuronavigation (mean failed antidepressants = 4.9 ± 3.0), although this difference did not reach significance (*p*= 0.30).

An additional subgroup analysis further subdivided this cohort into 63 patients who received left DLPFC treatment and who did not have any comorbidities. In this light, comorbidity was removed as a predictor variable. The average age of this group was 45.0 years (SD= 18.5) and included 39 men, 23 women, and 1 transgender person. In line with the previous analysis, the use of resting-state fMRI-guided targets (Estimate= 0.627, SE= 0.171, t= 3.669, *p*= 0.0007) was related to improved treatment outcomes, with an odds ratio of 1.87. Gender (Estimate= -0.276, SE= 0.124, t= -2.230, *p*= 0.032) also remained a predictor of better treatment outcome.

## DISCUSSION

The goal of the current study was to evaluate the real-world outcomes of accelerated rTMS treatment in an outpatient clinical setting, and identify key predictors of treatment response. Our findings highlight that accelerated rTMS represents a promising therapeutic option for individuals with treatment-resistant depression. Particularly, its effectiveness across a broad range of symptom profiles suggests that it can be tailored to meet the diverse needs of patients who have not responded to other depression treatments including pharmacotherapy, ECT, or conventional rTMS.

Our findings also highlight the value of resting-state fMRI targeted treatment approaches and patient-specific factors in managing treatment-resistant depression. Treatments guided by resting-state fMRI led to significantly better outcomes compared to scalp-based targeting, underscoring the potential of precision medicine to enhance treatment efficacy. Additionally, our analysis showed that women experienced more favorable outcomes in line with previous literature,^17,18^ suggesting a need to consider gender differences in treatment planning. Patients with comorbidities also had worse, though still good, outcomes with our current approach. This may be because we do not yet understand how to optimally neuro-navigate with more complex patients. The magnitude of the effect is grossly consistent with other estimates^17,18^.

Despite exploring variables traditionally linked to high treatment resistance, such past ECT and number of failed antidepressant trials, these factors did not significantly predict treatment outcomes. This finding suggests that accelerated rTMS may still be effective for patients who have faced multiple treatment failures.

While we found that fMRI targeting may provide added benefit, accelerated non-navigated treatments were also quite effective. For patients without access to neuro-navigated treatments, accelerated non-navigated treatments are a reasonable option.

These real-world data provide further evidence supporting the high response and remission rates observed with fMRI-guided rTMS for treatment-resistant depression in clinical trials^11^ and emphasize the importance of refining treatment protocols to better meet individual patient needs. Future prospective research to further explore the clinical potential of personalized rTMS approaches and to confirm these findings in larger, more diverse patient populations is warranted.

### Limitations

There are several limitations that should be acknowledged. First, the retrospective nature of the analysis may limit the ability to establish causality between treatment variables and outcomes. There may be unmeasured confounders that contribute to the observed significant effect of neuronavigation, although this concern is mitigated by the fact that patients receiving neuronavigation tended to be more treatment resistant. Second, the sample size, while adequate, may not fully capture the variability and potential outliers present in a broader population. In our clinical context, there is treatment selection bias as the cost to do neuronavigation is higher than non-navigation; we cannot rule out that accelerated TMS in general may be more effective for the very rich than the middle class. It may also be that there are time-based effects, with later patients being different than early patients. Additionally, the reliance on medical record data may introduce biases related to the accuracy and completeness of the records. Lastly, the study was conducted in a single outpatient clinical setting, which may limit the generalizability of the findings to other settings or populations.

## Conclusions

This study underscores the effectiveness of accelerated rTMS in treating treatment-resistant depression in a real-world outpatient setting, with fMRI-guided targeting and gender emerging as significant predictors of treatment response. The findings advocate for the incorporation of precision medicine techniques in the treatment of depression, particularly in patients with a history of treatment resistance. Future research should aim to overcome the limitations of this study through prospective, randomized trials to further validate and refine the use of accelerated rTMS, enhancing its efficacy and accessibility in clinical practice.

## Data Availability

Data produced in the present study may be available upon reasonable request to the authors.

## ACKNOWLEDGEMENTS

We thank Teresa Nguyen, Karen Marinov, Tonie Nguyen, Denise Rizvi, Joyce Pi, Shruthi Shankar, Julia Marco, Angel Luna, Justin Cheng, Gloria Pai, and Peter Sedaros for their contributions to the care and coordination of these patients.

